# Evaluation of bivalent Omicron BA.1 booster vaccination after different priming regimens in healthcare workers (SWITCH ON): a randomized controlled trial

**DOI:** 10.1101/2022.12.18.22283593

**Authors:** Ngoc H. Tan, Daryl Geers, Roos S.G. Sablerolles, Wim J.R. Rietdijk, Abraham Goorhuis, Douwe F. Postma, Leo G. Visser, Susanne Bogers, Laura L.A. van Dijk, Lennert Gommers, Leanne P.M. van Leeuwen, Annemarie Boerma, Sander H. Nijhof, Karel A. van Dort, Marion P.G. Koopmans, Virgil A.S.H. Dalm, Melvin Lafeber, Neeltje A. Kootstra, Anke L.W. Huckriede, Debbie van Baarle, Luca M. Zaeck, Corine H. GeurtsvanKessel, Rory D. de Vries, P. Hugo M. van der Kuy, the SWITCH Research Group

## Abstract

**Summary:** 

**Background:** Bivalent mRNA-based COVID-19 vaccines encoding the ancestral and Omicron spike protein were developed as a countermeasure against antigenically distinct SARS-CoV-2 variants. We compared the (variant-specific) immunogenicity and reactogenicity of mRNA-based bivalent Omicron BA.1 vaccines in individuals who were primed with adenovirus- or mRNA-based vaccines.

**Methods:** In this open-label, multicenter, randomized, controlled trial, healthcare workers primed with Ad26.COV2.S or mRNA-based vaccines were boosted with mRNA-1273.214 or BNT162b2 OMI BA.1. The primary endpoint was the fold change in S1-specific IgG antibodies pre- and 28 days after booster vaccination. Secondary outcomes were fast response, (antibody levels on day 7), reactogenicity, neutralization of circulating variants and (cross-reactive) SARS-CoV-2-specific T-cell responses.

**Findings:** No effect of different priming regimens was observed on bivalent vaccination boosted S1-specific IgG antibodies. The largest increase in S1-specific IgG antibodies occurred between day 0 and 7 after bivalent booster. Neutralizing antibodies targeting the variants in the bivalent vaccine (ancestral SARS-CoV-2 and Omicron BA.1) were boosted. In addition, neutralizing antibodies against the circulating Omicron BA.5 variant increased after BA.1 bivalent booster. T-cell responses were boosted and retained reactivity with variants from the Omicron sub-lineage.

**Interpretation:** Bivalent booster vaccination with mRNA-1273.214 or BNT162b2 OMI BA.1 resulted in a rapid recall of humoral and cellular immune responses independent of the initial priming regimen. Although no preferential boosting of variant-specific responses was observed, the induced antibodies and T-cells cross-reacted with Omicron BA.1 and BA.5. It remains crucial to monitor immunity at the population level, and simultaneously antigenic drift at the virus level, to determine the necessity (and timing) of COVID-19 booster vaccinations.

**Funding:** The Netherlands Organization for Health Research and Development (ZonMw) grant agreement 10430072110001.

**Research in context:** *Evidence before this study:* Vaccination against coronavirus disease-2019 (COVID-19) initially provided high levels of protection from both infection and severe disease. However, the emergence of antigenically distinct variants resulted in frequent breakthrough infections, especially with the emergence of variants from the Omicron sub-lineages. The frequent mutations in the Spike protein, and specifically the receptor binding domain (RBD), resulted in the recommendation by the WHO advisory group to update vaccines with novel antigens. Bivalent mRNA-based vaccines, encoding the Spike protein from both the ancestral SARS-CoV-2 and Omicron BA.1 (and later on BA.5) were subsequently introduced. Initial small comparative studies have been released on the evaluation of these bivalent vaccines, but it is essential is to evaluate the immunogenicity and reactogenicity of the vaccines against the background of different priming regimens.

*Added value of this study:* The SWITCH ON trial evaluated the bivalent booster vaccines BNT162b2 OMI BA.1 and mRNA-1273.214 vaccine in a cohort of Dutch healthcare workers. Study participants were primed with either Ad26.COV2.S, mRNA-1273, or BNT162b2. The study investigated three important topics: (1) immunogenicity of Omicron BA.1 bivalent vaccines after Ad26.COV2.S- or mRNA-based vaccine priming, (2) rapid immunological recall responses, indicative of preserved humoral and cellular immunological memory, and (3) cross-reactivity with relevant variants after booster vaccination.

*Implication of all the available evidence:* Vaccination with the bivalent booster mRNA-1273.214 or BNT162b2 OMI BA.1 resulted in a rapid recall of humoral and cellular immune responses independent of the initial priming regimen. The largest fraction of (neutralizing) antibodies and virus-specific T-cells was recalled within 7 days post booster vaccination. Although no preferential boosting of variant-specific responses was observed, the induced antibodies and T-cells cross-reacted with Omicron BA.1, which was included in the vaccine, but also the more antigenically distinct BA.5. It remains crucial to monitor immunity at the population level, and simultaneously antigenic drift at the virus level, to determine the necessity (and timing) of COVID-19 booster vaccinations.

## Introduction

The ongoing spread of severe acute respiratory syndrome coronavirus-2 (SARS-CoV-2) remains a public health emergency of international concern. Although vaccination against coronavirus disease-2019 (COVID-19) initially provided high levels of protection from both infection and severe disease^1,2^, emergence of variants has resulted in escape from protection against infection. Frequent breakthrough infections can be explained by a combination of waning antibodies in combination with (partial) evasion from neutralizing antibodies, especially since the emergence of the Omicron sublineages^3-7^. The shift in key epitopes, which were characteristic of Omicron variant viruses compared to ancestral viruses^8^, resulted in the recommendation by the WHO advisory group on vaccinations to update the vaccines^9^. This led to the introduction of bivalent spike (S) vaccines.

The first generation of licensed bivalent vaccines consisted of mRNAs encoding the S protein from both the ancestral SARS-CoV-2 and Omicron BA.1^10,11^. A study with the bivalent mRNA-1273.214 vaccine induced higher levels of Omicron BA.1 neutralizing antibodies compared to the monovalent mRNA-1273 vaccine, when given as a second booster in adults who had previously received a primary vaccination series and first booster with the mRNA-1273 vaccine^10^. Partially based on this result, many countries introduced bivalent vaccines in their booster campaigns.

We have now entered a phase in the pandemic in which (1) repeated boosters are available for risk groups and the general population, (2) a relatively low number of severe COVID-19 cases and low mortality rate are observed, but (3) SARS-CoV-2 continues to display antigenic drift^8^. By the time updated bivalent vaccines were introduced, further diverged Omicron sub-lineages had become dominant. To ascertain the immunological benefit of additional (bivalent) booster vaccinations and provide scientific evidence for decision makers, in-depth evaluations of immunogenicity are important.

The SWITCH ON trial^12^ aims to evaluate the bivalent booster vaccines BNT162b2 OMI BA.1 and mRNA-1273.214 vaccine in a cohort of Dutch healthcare workers. The study investigates three main topics: (1) immunogenicity of Omicron BA.1 bivalent vaccines after Ad26.COV2.S- or mRNA-based vaccine priming, (2) rapid immunological recall responses, indicative of preserved humoral and cellular immunological memory, and (3) cross-reactivity with relevant variants after booster vaccination. These data will aid in the discussion on the necessity of future booster vaccinations in the healthy general population and aim to facilitate more personalized vaccination approaches in future public health interventions against COVID-19.

## Methods

### Trial oversight

The SWITCH ON study is an open-label, multicenter, randomized, controlled trial involving healthcare workers from four academic hospitals in the Netherlands^12^. The trial protocol was approved by the medical ethics committee of Erasmus Medical Centre, the sponsor site, and the local review boards of the other participating centers. The study adheres to the principles of the Declaration of Helsinki. All eligible subjects provided written informed consent before their participation in the study.

### Trial design

Healthcare workers were eligible to participate in the SWITCH ON study if they were between the age of 18 and 65, and completed a primary vaccination regimen with either Ad26.COV2.S (1 shot) or an mRNA-based (BNT162b2 or mRNA-1273, 2 shots) vaccine. All participants had also received at least one booster dose with an mRNA-based vaccine, given no later than three months before the start of the SWITCH ON study. Participants with severe comorbidities such as dialysis dependence, or participants with an immunodeficiency due to treatment with immunosuppressants or cancer therapy were excluded from the study. Participants with a known history of prior SARS-CoV-2 infection were eligible, unless the infection occurred less than three months before the start of the study (based on self-reporting). At baseline, SARS-CoV-2 nucleocapsid (N)-specific antibodies were measured to determine the distribution of infection history across groups. Participants with positive nucleocapsid test, who did not report having contracted SARS-CoV-2 less than three months before the start of the study, were included in the statistical analyses. The full list of in- and exclusion criteria was published previously^12^. The representativeness of the study participants is described in **Table S1**.

Block randomization was used to randomize participants equally into a direct boost (DB) group or a postponed boost (PPB) group, with stratification for Ad26.COV2.S-priming or mRNA-based priming. Participants in the DB group received the Omicron BA.1 bivalent booster vaccine in October 2022, whereas participants in the PPB group will receive a bivalent booster vaccine in December 2022. Participants of 45 years of age or older were administered mRNA-1273.214 (50μg) and participants younger than 45 years of age received BNT162b2 OMI BA.1 (30μg), following the guidelines of the Dutch National Institute for Public Health and the Environment (RIVM)^13^. Based on their respective priming regimens, the participants of the DB group were further divided into four subgroups: (i) Ad26.COV2.S prime and mRNA-1273.214 boost (Ad/M), (ii) Ad26.COV2.S prime and BNT162b2 OMI BA.1 boost (Ad/P), (iii) mRNA-based prime and mRNA-1273.214 boost (mRNA/M), and (iv) mRNA-based prime and BNT162b2 OMI BA.1 boost (mRNA/P). Blood samples were collected on the day of booster vaccination (day 0, study visit 1, SV1), and 7 (SV2) and 28 days (SV3) after booster (**Figure S1)**. Additionally, 25% of the samples in each arm were randomly selected for in-depth immunological analyses.

### Immunogenicity

SARS-CoV-2-specific antibodies and T-cell responses were measured in all participants at baseline, and 7 and 28 days after booster vaccination. Ancestral S1-specific IgG antibody levels were measured using the Liaison SARS-CoV-2 TrimericS IgG assay (DiaSorin) as previously described^4^. S-specific binding antibodies to the ancestral and Omicron BA.1 and BA.5 S proteins were assessed by ELISA as previously described^14,15^. Neutralizing antibodies targeting the ancestral and Omicron sub-lineages (BA.1 and BA.5) were assessed by plaque reduction neutralization test (PRNT) as previously described^14,16^. SARS-CoV-2-specific T-cell responses were measured by interferon-γ release assay (IGRA) using the QuantiFERON SARS-CoV-2 kit (QIAGEN)^3,4^. Additionally, SARS-CoV-2-specific T-cells were phenotyped in an activation-induced marker (AIM) flow cytometry assay^3^. In these AIM assays, cross-reactive variant-specific T-cell responses targeting the ancestral, Omicron BA.1 and Omicron BA.5 variants were measured^3,14,15^.

### Reactogenicity

Participants received an electronic questionnaire on day 8 after booster, inquiring about the adverse reactions^4^. The severity of adverse reactions was described in accordance with the Toxicity Grading Scale for Healthy Adult and Adolescent Volunteers Enrolled in Preventive Vaccine Clinical Trials^17^. Any serious adverse reactions, both local and systemic, were additionally reported via email and phone calls.

### Statistical analysis

We calculated that the study required 91 participants in each of four arms (Ad/M, Ad/P, mRNA/M, mRNA/P) to achieve 80% power at two-sided 5% significance level to detect a log10 transform difference of 0.2 in fold change 28 days after booster vaccination. Considering a 10% loss-to-follow-up margin, we included 100 participants per arm, resulting in a total sample size of 400 participants. These consisted of 200 participants primed with Ad26.COV2.S and 200 participants primed with BNT162b2 or mRNA-1273, equally divided over the DB and PPB groups. During study recruitment, a new COVID-19 booster policy by the national government of the Netherlands^13^ was implemented with the introduction of different vaccine strategies for the 2 available vaccines according to age (above and below 45 years), which was not part of the initial power calculation.

Descriptive analysis was used to report baseline characteristics of participants. For continuous variables with normal distribution, median and interquartile range (IQR) were reported. Categorical variables were presented as numbers (percentage). For the immunogenicity data we also present the geometric mean titer values (GMT).

For the primary outcome, the fold change antibody response (day 0 versus day 28) was compared between the Ad26.COV2.S-primed and mRNA-based-primed (Ad/P vs. mRNA/P and Ad/M vs. mRNA/M) using a Mann Whitney U test. The pre-specified secondary endpoints were fast response, levels of neutralizing antibodies, S-specific T-cell responses and reactogenicity. A fast response was defined as having reached an immune response on day 7 that is equal to or higher than 65% increase of the titer on day 28 post-vaccination. The fast response was reported per priming regimen and booster vaccination as percentage. We reported SARS-CoV-2-specific T-cell responses and neutralizing antibody titers using similar statistics as in analyses for the primary endpoint. Post-hoc analyses of S-specific binding antibodies measured by ELISA and S-specific cross-reactive T-cell responses measured by AIM provided an overview of the variant-specific immunogenicity following bivalent vaccination. We used p<0.01 as the statistical significance threshold. We reported missing values when applicable (**Table S4**), and as number of missing values are low no imputation is used. All statistical analyses were conducted using GraphPad Prism software (version 9.4.1) and Rstudio (version 4.2.1). The study is registered with ClinicalTrials.gov, NCT05471440.

## Results

### Baseline characteristics of the participants

Of the 592 healthcare workers who were screened for eligibility, 434 were included into the SWITCH ON study. Excluded participants did not meet the inclusion criteria (N=62), withdrew before randomization (N=8) or were excluded for logistical reasons (N=88). 219 participants were randomized into the DB group for which the results will be discussed in this paper. Thirty-two participants were excluded from the intention-to-treat analysis because of withdrawal from the study, SARS-CoV-2 breakthrough infection in between study visits (SV), or missing blood samples across study visits (**Figure 1**). All included participants in the intention-to-treat analyses (N=187) adhered to the defined timing intervals in-between study visits. The median interval between SV1 and SV2 was 7.0 days (IQR, 7.0-7.0) and 28.0 days (IQR, 28.0-28.0) between SV1 and SV3. The median interval between the last vaccination and the bivalent booster vaccination was 298 days (IQR, 266.0-309.5). Notably, the distribution of sex was different between study groups with a higher number of female participants across all groups. Recent SARS-CoV-2 breakthrough infections were most frequently reported in the mRNA/M group, as assessed by N-specific antibody levels in serum at baseline. **Table 1** presents the complete baseline characteristics.

**Figure 1.**
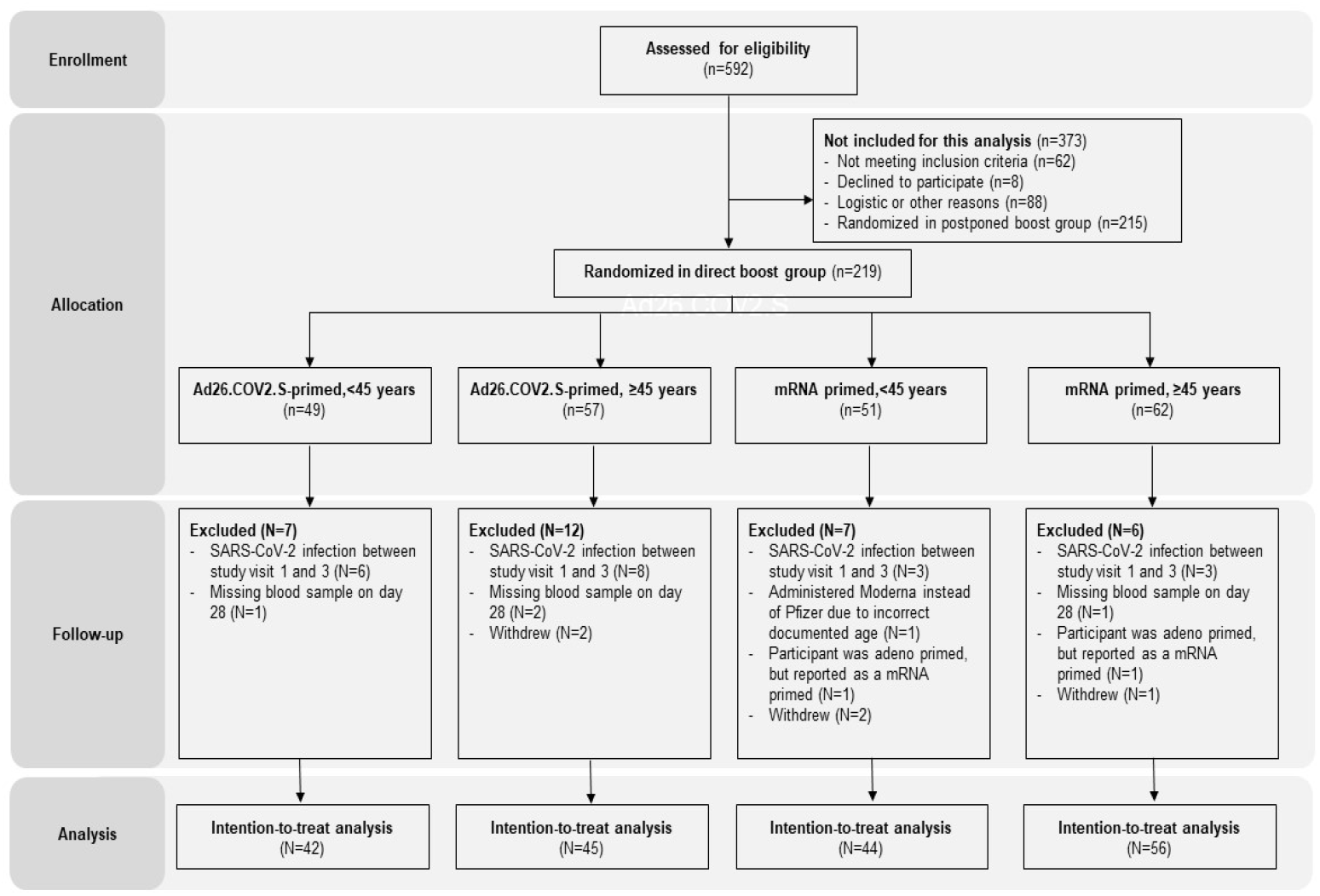
Participant screening, inclusion, exclusion, and analysis. 592 healthcare workers (HCW) were screened for eligibility, of whom 62 did not meet inclusion criteria, 8 withdrew from the study before randomization, and 88 were excluded for logistical reasons. Ultimately, 434 HCW were included into the study and randomized into a direct boost (DB; n=219) or postponed boost (PPB; n=215) group (further explained in **Figure S1**). Participants were allocated into 1 of 4 study arms. For final analysis, several participants were excluded because of SARS-CoV-2 breakthrough infection in between study visits, missing blood samples, or withdrawal from the study, leading to a final inclusion of n=187 participants in an intention-to-treat analysis.

**Table 1.** Baseline characteristics of trial participants

|  |  | Total<br>N = 187 | Ad26.COV2.S primed,<br><45 years (Ad/P)<br>N = 42 | mRNA primed,<br><45 years<br>(mRNA/P)<br>N = 44 | Ad26.COV2.S<br>primed, ≥45 years<br>(Ad/M)<br>N = 45 | mRNA primed,<br>≥45 years (mRNA/M)<br>N = 56 |
| --- | --- | --- | --- | --- | --- | --- |
| Sex | Male | 49 (26%) | 15 (36%) | 10 (23%) | 17 (38%) | 7 (13%) |
|  | Female | 138 (74%) | 27 (64%) | 34 (77%) | 28 (62%) | 49 (88%) |
|  | Age (years) | 47.0 (36.0 - 53.0) | 32.0 (28.0-40.5) | 35.0 (28.0-40.0) | 52.0 (50.0 - 55.0) | 53.0 (50.0 - 57.0) |
|  | BMI (kg/m<sup>2</sup>) | 24.2 (22.5 - 27.0) | 23.8 (22.2-24.8) | 23.8 (21.7 - 26.3) | 25.0 (23.0-27.5) | 24.8 (23.0 - 27.7) |
| Ancestry | African | 2 (1%) | 0 (0%) | 1 (2%) | 1 (2%) | 0 (0%) |
|  | Asian | 6 (3%) | 0 (0%) | 3 (7%) | 0 (0%) | 3 (5%) |
|  | European | 173 (93%) | 40 (95%) | 39 (89%) | 44 (98%) | 50 (89%) |
|  | North American | 1 (1%) | 0 (0%) | 1 (2%) | 0 (0%) | 0 (0%) |
|  | South American | 0 (0%) | 0 (0%) | 0 (0%) | 0 (0%) | 0 (0%) |
|  | Other | 5 (3%) | 2 (5%) | 0 (0%) | 0 (0%) | 3 (5%) |
| Occupation in<br>hospital | Administrative/policy maker | 27 (14%) | 6 (14%) | 3 (7%) | 10 (22%) | 8 (14%) |
|  | Medical doctor | 15 (8%) | 2 (5%) | 4 (9%) | 0 (0%) | 9 (16%) |
|  | Facility services | 3 (2%) | 1 (2%) | 1 (2%) | 1 (2%) | 0 (0%) |
|  | Management | 24 (13%) | 5 (12%) | 3 (7%) | 9 (20%) | 7 (13%) |
|  | Supportive staff clinic/emergency<br>department | 2 (1%) | 1 (2%) | 0 (0%) | 0 (0%) | 1 (2%) |
|  | Supportive staff outpatient clinic | 1 (1%) | 0 (0%) | 0 (0%) | 0 (0%) | 1 (2%) |
|  | Researcher | 45 (24%) | 17 (40%) | 15 (34%) | 5 (11%) | 8 (14%) |
|  | Nurse | 11 (6%) | 0 (0%) | 4 (9%) | 0 (0%) | 7 (13%) |
|  | Other | 59 (32%) | 10 (24%) | 14 (32%) | 20 (44%) | 15 (27%) |
| Comorbidities | Cardiovascular diseases | 2 (1%) | 0 (0%) | 0 (0%) | 0 (0%) | 2 (4%) |
|  | Pulmonary diseases | 9 (5%) | 1 (2%) | 2 (5%) | 2 (4%) | 4 (7%) |
|  | Diabetes mellitus | 3 (2%) | 0 (0%) | 1 (2%) | 2 (4%) | 0 (0%) |
|  | Liver diseases | 2 (1%) | 0 (0%) | 0 (0%) | 0 (0%) | 2 (4%) |
|  | Kidney diseases | 3 (2%) | 1 (2%) | 1 (2%) | 0 (0%) | 1 (2%) |
| S1-specific<br>binding antibodies<br>(LIAISON,<br>BAU/ml) | Study visit 1 | 2660 (1530-6920) | 1950 (1252.5-3422.5) | 2725 (1560-7927.5) | 2130 (1210-4760) | 5665 (2007.5-11050) |
| Interferon gamma<br>release assay<br>(IGRA, IU/ml) | Study visit 1 | 0.36 (0.14-0.76) | 0.47 (0.15-0.64) | 0.53 (0.19-1.06) | 0.19 (0.13-0.63) | 0.46 (0.13-0.94) |
| Nucleocapsid | Negative | 151 (81%) | 33 (79%) | 39 (89%) | 41 (91%) | 38 (68%) |
|  | Positive | 36 (19%) | 9 (21%) | 5 (11%) | 4 (9%) | 18 (32%) |
| Original center | Amsterdam University Medical<br>Centers | 22 (12%) | 6 (14%) | 8 (18%) | 3 (7%) | 5 (9%) |
|  | Erasmus Medical Center, Rotterdam | 130 (70%) | 23 (55%) | 34 (77%) | 23 (51%) | 50 (89%) |
|  | Leiden University Medical Center | 16 (9%) | 9 (21%) | 0 (0%) | 7 (16%) | 0 (0%) |
|  | University Medical Center Groningen | 19 (10%) | 4 (10%) | 2 (5%) | 12 (27%) | 1 (2%) |
| Median time | Between study 1 and 2 (day 0 and 7) | 7.0 (7.0 - 7.0) | 7.0 (7.0 - 7.0) | 7.0 (7.0 - 7.0) | 7.0 (7.0 - 7.0) | 7.0 (7.0 - 7.0) |
|  | Between study 1 and 3 (day 0 and 28) | 28.0 (28.0 - 28.0) | 28.0 (28.0 - 28.0) | 28.0 (28.0 - 28.0) | 28.0 (28.0 - 28.0) | 28.0 (28.0 - 28.0) |
|  | Between last booster and bivalent booster (in days) | 298.0 (266.0-309.5) | 266.5 (260.5-305.3) | 303.0 (297.0-309.3) | 266.0 (263.0-293.0) | 307.0 (302.3-310.3) |

*Note: For categorical variables we present numbers (percentages), whereas continuous variables we present median (interquartile range).*

### S1-specific binding antibodies after a bivalent booster

S1-specific binding antibody levels increased 7 days after bivalent booster and remained stable over 28 days in all four study groups (**Figure 2A)**. No significant difference was observed between the Ad/P (2.9-fold) and mRNA/P (3.6-fold) groups (p=0.12), whereas a trend (p=0.03) towards increased fold changes after bivalent booster vaccination was observed for the mRNA/M (3.5-fold) and Ad/M (4.3-fold) groups. However, higher baseline levels of S1-specific binding IgG antibodies were detected in mRNA-primed individuals (geometric mean titer (GMT), mRNA/M 5,196 and mRNA/P 3,198 BAU/mL) than in Ad26.COV2.S-primed individuals (GMT, Ad/M 2,032 and Ad/P 1,959 BAU/mL) (**Figure 2A**). Similarly, S1-specific binding IgG antibody levels were higher in the mRNA/M (GMT, 18,329 BAU/mL) and mRNA/P (GMT, 11,643 BAU/mL) study groups compared to the Ad/M (GMT, 8,685 BAU/mL) and Ad/P (GMT, 5,740 BAU/mL) study groups 28 days after bivalent booster vaccination (**Figure 2A)**.

**Figure 2.**
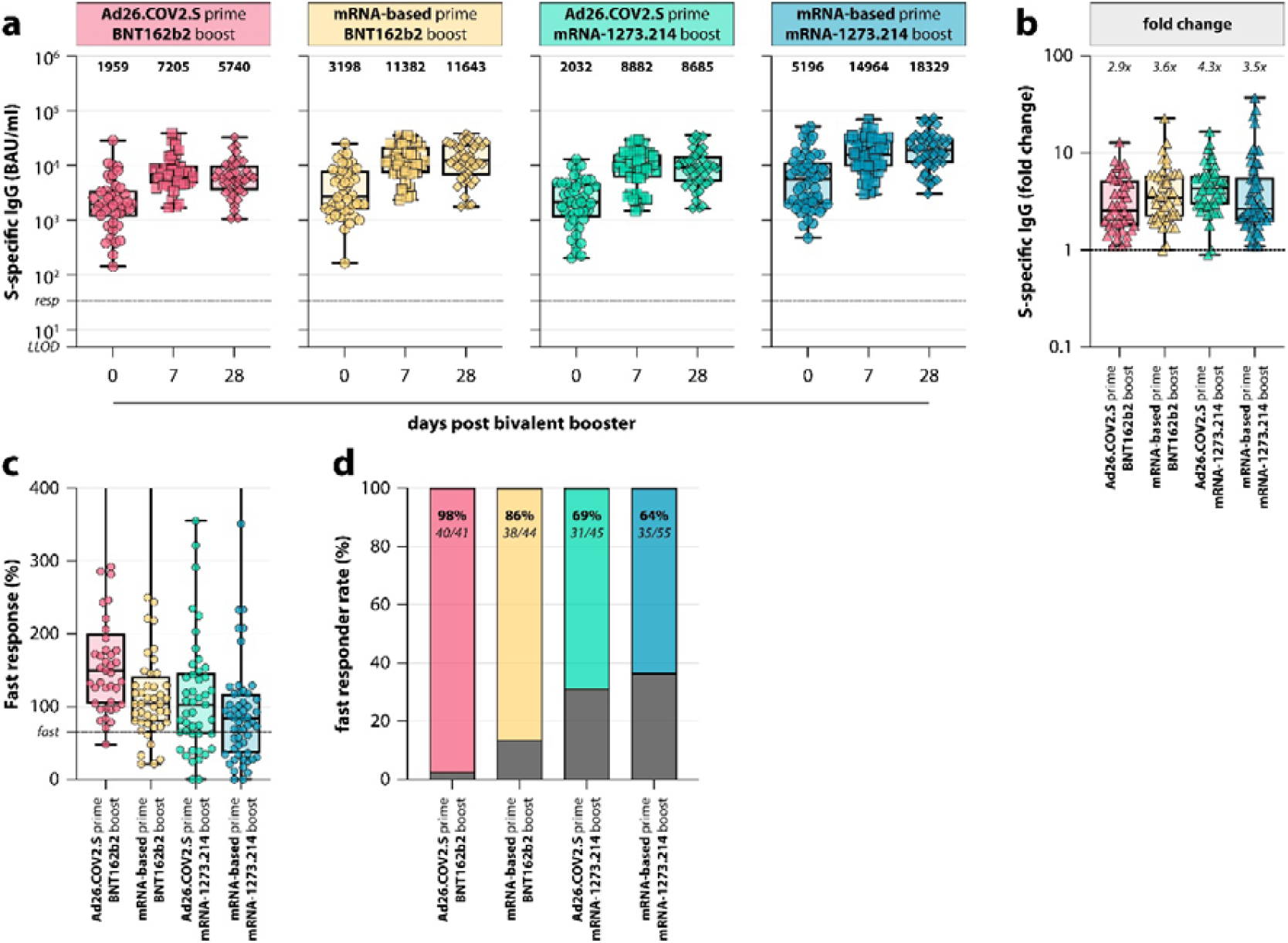
S-specific binding antibodies and fast response after bivalent booster vaccination. **(a)** Detection of (ancestral) S1-specific binding IgG antibodies at baseline before bivalent booster (0 days; circles), and 7 days (squares) and 28 days (diamonds) after booster in the four study groups (Ad26.COV2.S prime / BNT162b2 OMI BA.1 boost, red, Ad/P; mRNA-based prime / BNT162b2 OMI BA.1 boost, yellow, mRNA/P; Ad26.COV2.S prime / mRNA-1273.214 boost, teal, Ad/M; mRNA-based prime / mRNA-1273.214 boost, blue, mRNA/M). The lower limit of detection (LLOD) was set at 4.81 binding antibody units per milliliter (BAU/mL). The cut-off responder value was set at 33.8 BAU/mL (horizontal dashed line). The bold numbers above the plots indicate the geometric mean titer (GMT) per time point. **(b)** Fold change of S-specific binding antibody levels between baseline and 28 days after bivalent booster vaccination in the four study groups. The horizontal dashed line indicates a fold change of 1, which corresponds to no increase or decrease in S1-specific binding antibody levels post-vaccination. Exact fold changes per study group are indicated above the respective plot. **(c)** Fast response based on S1-specific antibody levels in the four study groups. The horizontal dashed line indicates the fast responder cut-off, which was defined as having reached an antibody level on day 7 post-vaccination that is equal to or higher than 65% of the total S-specific binding antibody response on day 28. **(d)** Percentage of fast responders per study group.

### Rapid recall of S1-specific antibodies after a bivalent booster

As a secondary outcome, the fast response of S1-specific antibodies following bivalent booster vaccine was calculated. The largest increase in S1-specific binding IgG antibodies occurred between day 0 and day 7 after booster in all four study groups **(Figure 2C and 2D)**. Notably, the proportion of fast responders for participants who received the BNT162b2 OMI BA.1 bivalent booster vaccination was higher (Ad/P: 98%; mRNA/P: 86%) in comparison with those having received the mRNA-1273.214 bivalent vaccine (Ad/M: 69%; mRNA/M: 64%) (**Figure 2D**). Two participants from the total of 187 were excluded from the analysis due to missing SV2 blood draw.

### Variant-specific antibodies after a bivalent booster

To assess boosting of SARS-CoV-2-CoV-2-neutralizing antibodies after bivalent booster vaccination, plaque reduction neutralization tests (PRNT) using ancestral SARS-CoV-2, and the Omicron BA.1 and BA.5 variants were performed on a random selection of serum samples (**Figure 3, Figure S2**). Neutralizing antibody levels against ancestral SARS-CoV-2 were comparable at baseline across all four study groups, although participants in the mRNA/M group had higher neutralizing antibody titers (**Figure 3D**). Bivalent booster vaccination increased the neutralizing antibody titers against ancestral SARS-CoV-2 in all study groups at 7 and 28 days post-vaccination. Similar to the binding antibodies (**Figure 2**), the largest increase in neutralizing antibodies was observed between day 0 and day 7 (**Figure 3**). At baseline, the neutralizing antibody response of the participants against Omicron BA.1 and BA.5 variants was markedly lower compared to neutralizing antibody levels against ancestral SARS-CoV-2. Neutralizing antibody levels against BA.1 and BA.5 increased after bivalent booster vaccination with a similar pattern of increase compared to antibodies neutralizing the ancestral SARS-CoV-2; the magnitude of boosting was comparable between the different priming regimens and booster vaccinations at all three study visits.

**Figure 3.**
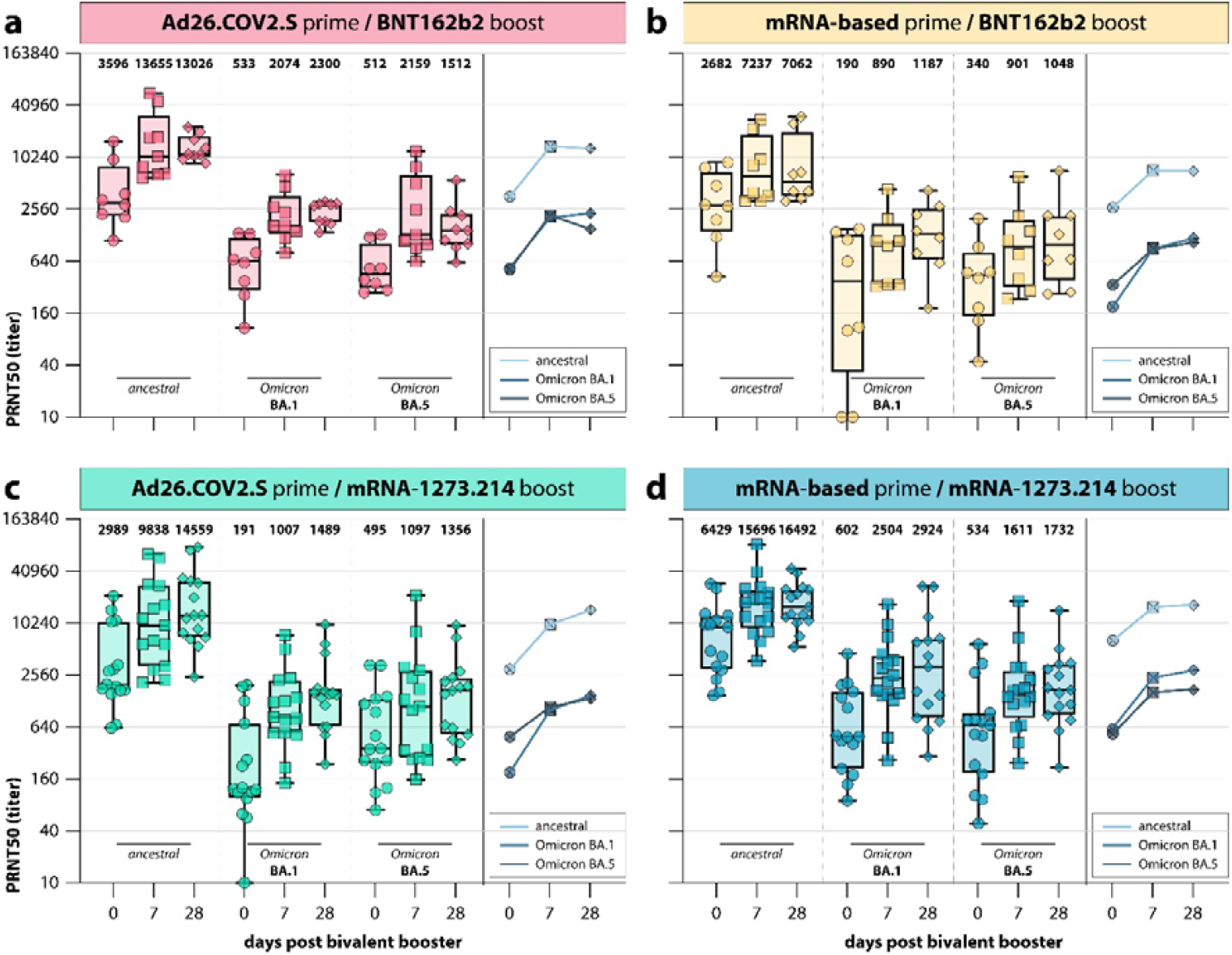
Variant-specific neutralization after bivalent booster vaccination. **(a-d)** Detection of SARS-CoV-2-neutralizing antibodies targeting ancestral SARS-CoV-2 and Omicron variants BA.1 and BA.5 at baseline (0 days; circles), 7 days (squares), and 28 days (diamonds) after bivalent booster vaccination for **(a)** Ad26.COV2.S prime / BNT162b2 OMI BA.1 boost (red; Ad/P), **(b)** mRNA-based prime / BNT162b2 OMI BA.1 boost (yellow; mRNA/P), **(c)** Ad26.COV2.S prime / mRNA-1273.214 boost (teal; Ad/M), and **(d)** mRNA-based prime / mRNA-1273.214 boost (blue; mRNA/M). The bold numbers above the plots indicate the geometric mean titer (GMT) per time point and variant. The line graphs next to each panel represent the median neutralizing titer against ancestral SARS-CoV-2 and Omicron variants BA.1 and BA.5 at baseline (0 days), 7 days, and 28 days after bivalent booster vaccination. When no neutralization was observed, the PRNT50 was given a value of 10.

A similar pattern was observed for S-specific binding antibody levels against ancestral SARS-CoV-2, and the Omicron BA.1 and BA.5 variants when measured by ELISA (**Figure S3**). S-specific binding antibody levels for ancestral SARS-CoV-2, and Omicron BA.1 and BA.5 variants were comparable at baseline, increased 7 days after bivalent booster vaccination and remained stable over 28 days for all four study groups. S-specific binding antibody titers against Omicron BA.1 and BA.5 were significantly lower than against ancestral SARS-CoV-2 in all four study groups at all the time points (**Figure S3**).

### S-specific T-cell responses after a bivalent booster

SARS-CoV-2 S-specific T-cell responses were assessed by measuring IFN-γ levels after stimulating whole blood with S-specific peptide pools. T-cell responses increased directly after bivalent booster (day 7) and then decreased at day 28 post-booster vaccination for all four study groups (**Figure 4A, 4B, and Figure S4**). T-cell responses were comparable at baseline and after bivalent booster for all four study groups (**Figure 4A**). No difference in fold change of IFN-γ levels was observed between the BNT162b2 OMI BA.1 boosted groups (Ad/P: 2.8-fold and mRNA/P: 2.1-fold), however, a higher fold change in IFN-γ levels was observed after bivalent booster vaccination with mRNA-1273.214 following Ad26.COV2.S priming (Ad/M: 4.3-fold) compared to mRNA priming (mRNA/M: 2.3-fold) (**Figure 4B**).

**Figure 4.**
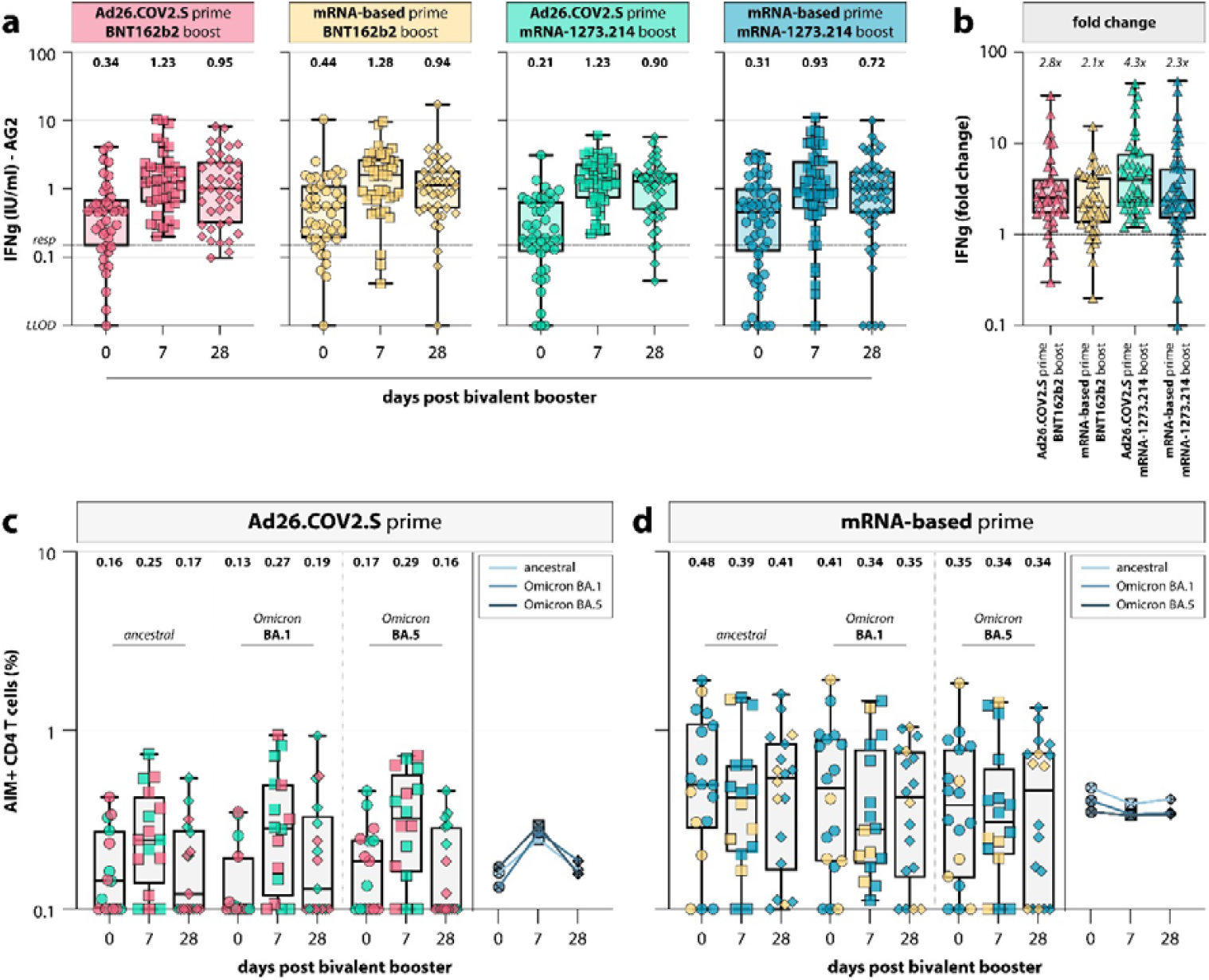
Variant-specific T-cell responses after bivalent booster vaccination. **(a)** Detection of IFN-γ in international units per milliliter (IU/mL) following stimulation of whole blood with overlapping S peptide pools in coated QuantiFERON tubes at baseline (day 0; circles), 7 days (squares), and 28 days (diamonds) after bivalent booster vaccination in the four study groups (Ad26.COV2.S prime / BNT162b2 OMI BA.1 boost, red, Ad/P; mRNA-based prime / BNT162b2 OMI BA.1 boost, yellow, mRNA/P; Ad26.COV2.S prime / mRNA-1273.214 boost, teal, Ad/M; mRNA-based prime / mRNA-1273.214 boost, blue, mRNA/M). A lower limit of detection (LLOD) was set at 0.01 IU/mL as per manufacturer’s instructions. The horizontal dotted line indicates a pre-defined responder cut-off of 0.15 IU/mL. The bold numbers above the plots indicate the geometric mean (GM) IFN-γ levels per timepoint. **(b)** Fold change of IFN-γ levels between baseline and 28 days after bivalent booster vaccination in the four study groups. The horizontal dashed line indicates a fold change of 1, which corresponds to no increase or decrease. Exact fold changes per study group are indicated above the respective plot. **(c, d)** Percentages of activation-induced marker (AIM)-positive CD4+ T-cells following *ex vivo* stimulation with an overlapping peptide pool spanning the full S protein of ancestral SARS-CoV-2 or the Omicron variants BA.1 and BA.5 in **(c)** Ad26.COV2.S primed or **(d)** mRNA primed individuals. The symbol colors refer to their respective study group. The bold numbers above the plots indicate the geometric mean (GM) of the percentage of AIM-positive CD4+ T-cells per time point. The line graphs next to each panel represent the median percentage of AIM-positive CD4+ T-cells for ancestral SARS-CoV-2 and Omicron variants BA.1 and BA.5 at baseline (0 days), 7 days, and 28 days after bivalent booster vaccination.

S-specific T-cell cross-reactivity to BA.1 and BA.5 was assessed by AIM flow cytometry assay, after stimulating peripheral blood mononuclear cells (PBMC) with overlapping S peptides. SARS-CoV-2 specific CD4+ T-cell responses were generally higher in mRNA-primed individuals than in Ad26.COV2.S-primed individuals at baseline (geometric mean (GM), mRNA prime: 0.48 and Ad26.COV2.S prime: 0.16), 7 days (GM, mRNA prime: 0.39 and Ad26.COV2.S prime: 0.25), and 28 days after bivalent booster vaccination (GM, mRNA prime: 0.41 and Ad26.COV2.S prime: 0.17) (**Figure 4C and 4D**). An increase in S-specific T-cells between 0 and 7 days post vaccination was observed in the Ad26.COV2.2-primed participants, but not in the mRNA-based primed participants. Notably, CD4+ T-cells were cross-reactive with the Omicron BA.1 and BA.5 variants in all study groups (**Figure 4C and 4D**). CD8+ T-cell responses followed similar kinetics as CD4+ T-cell responses for all study groups (**Figure S5**).

### Reactogenicity

The reactogenicity data showed that pain at the injection site, muscle aches, headache or fatigue were the most reported side effects in all four study groups. All side effects were mild to moderate in severity (**Table S2**) and resolved within 48 hours (**Table S3**). The severity of side effects was similar between all groups, except for joint pain, which was experienced more frequently in mRNA-1273.214-boosted participants regardless of their priming regimen.

## Discussion

This study reports the immunogenicity and reactogenicity of BA.1 bivalent COVID-19 booster vaccines in Dutch healthcare workers primed with Ad26.COV2.S, mRNA-1273, or BNT16b2 vaccines. Bivalent booster vaccination significantly increased binding and neutralizing antibody levels in all groups and no differences in ‘boostability’ were observed between Ad26.COV2.S- and mRNA-based vaccine primed individuals.

We observed a clinical significant fold change in antibody titers from day 0 to day 28 in the four groups, though no differences between various priming regimes. Further, the largest increase in antibody titers within the first 7 days after booster vaccination. This was most prominent in BNT162b2 OMI BA.1 boosted groups. As per Dutch policy, those under 45 years of age received BNT162b2 OMI BA.1, whereas participants above 45 years were boosted with mRNA-1273.214, making it likely that the lower age of these participants is responsible for this difference. A similar rapid recall was noticed for T-cell responses measured in whole blood, with kinetics slightly different from the antibody kinetics: after the rapid recall at day 7, IFN-γ responses in whole blood slightly decreased at day 28 after booster vaccination. The observation of combined rapid SARS-CoV-2-specific antibody and T-cell recall responses is indicative of efficient induction of immunological memory by previous vaccinations and/or infections.

The BA.1 bivalent booster vaccination boosted neutralizing antibodies targeting both BA.1 and BA.5 from the Omicron sub-lineage, however, these neutralizing antibody levels were generally lower than those against ancestral SARS-CoV-2. This is consistent with previous studies describing the immunogenicity of BA.1 bivalent booster vaccines^10,18,19^. Although our data support the induction of cross-neutralizing antibodies by the BA.1 bivalent booster vaccines against emerging variants not contained in the vaccine (such as BA.5), we did not observe preferential boosting of BA.1 over BA.5 neutralizing antibodies. This is consistent with two recent studies that show that exposure to antigenically distinct Omicron variants^8^ by either vaccination or infection recalls pre-existing memory B-cells specific for epitopes shared by different SARS-CoV-2 variants^20,21^. Real-world data exploring the effectiveness of this increased breadth of the immune response will be essential when evaluating the need for continuous updating of variant-specific booster vaccines.

Whereas we did not observe any effect of the respective priming regimen on (neutralizing) antibodies before or after bivalent booster vaccination, this was different for CD4+ T-cell responses measured by AIM assay. Activation of CD4+ T-cells after stimulation with overlapping S peptide pools at any time point was considerably lower in Ad26.COV2.S-primed individuals compared to mRNA-primed individuals. This was similar to observations after primary vaccination^3,14^. Although the levels of SARS-CoV-2-specific CD4+ T cell levels in peripheral blood were lower in Ad26.COV2.S-primed participants at baseline, the response was rapidly reactivated upon antigen exposure. In all groups, T-cell responses generally displayed cross-reactivity with the Omicron BA.1 and BA.5 variants^3,14,22^, and no preferential induction of variant-specific T-cells was observed after booster vaccination.

Immunological memory is crucial in the prevention of severe COVID-19, as sterile immunity against SARS-CoV-2 infection cannot be achieved by vaccination or infection, which is illustrated by the low vaccine effectiveness against infection in HCW of 7-11%^23^. Our data show that bivalent booster vaccination leads to a robust recall of memory B- and T-cell responses, and that the largest fraction of these responses occurs within the first 7 days after boost. A similar rapid recall of immune memory is to be expected in the case of a SARS-CoV-2 breakthrough infection. It is therefore not unreasonable to assume that the generally mild disease profile upon infection with variants from the Omicron sub-lineage is at least partly driven by a broad memory recall response, which could be predictive of mild clinical disease after re-infection with future variants as well. This calls for a re-evaluation of the necessity and frequency of future COVID-19 booster vaccinations in the general population and risk groups. For specific populations, clinical evaluations should include the monitoring of immunity and severity of clinical disease, against the background of antigenic drift at the virus level.

## Data Availability

All data produced in the present study are available upon reasonable request to the authors.

## Author contributions

Conceptualization: P.H.M.v.d.K., C.G.v.K., R.D.d.V. Formal analysis: N.H.T., D.G., R.S.G.S., W.J.R.R., R.D.d.V. Funding acquisition: N.H.T., R.S.G.S, W.J.R.R., A.G., D.F.P., L.G.V., M.P.G.K., V.A.S.H.D., M.L., N.A.K., A.L.W.H., D.v.B., L.M.Z., C.G.v.K., R.D.d.V., P.H.M.v.d.K. Investigation: N.H.T., D.G., R.S.G.S, W.J.R.R., A.G., D.F.P., L.G.V., S.B., L.L.A.v.D., L.G., L.P.M.v.L., A.B., S.H.N., K.A.v.D., M.P.G.K., V.A.S.H.D., M.L., N.A.K., A.L.W.H., D.v.B., L.M.Z, C.G.v.K., R.D.d.V., P.H.M.v.d.K. Supervision: P.H.M.v.d.K., C.G.v.K., R.D.d.V. Visualization: D.G., L.M.Z., R.D.d.V. Writing-original draft: N.H.T., D.G., R.S.G.S., W.J.R.R., R.D.d.V., C.G.v.K., P.H.M.v.d.K. Writing: review and editing: all authors reviewed and edited the final version.

## Data sharing agreement

Anonymized individual participant data, analytics code, and other supporting documents will be made available when the study is complete, on reasonable requests made to the corresponding author.

## Acknowledgements

We acknowledge QIAGEN for supporting the study by providing QuantiFERON SARS-CoV-2 RUO Starter and Extended Packs. QIAGEN had no role in study design, data acquisition and analysis. Assay methodology images were created with Biorender.

